# An integrated AI-microfluidic platform reveals the broad persistence and developmental potential of rare sperm in non-obstructive azoospermia

**DOI:** 10.64898/2026.06.18.26355896

**Authors:** Huaying Chen, Peilin Chen, Weiqiang Xiao, Lei Wang, Mingzhe Song, Xiaoying Liu, Rui Shen, Sitong Guo, Jiaxi Li, Wentao Zhao, Meilan Mo, Chunyu Huang, Shiru Xu, Qing Sun, Huixian Zhong, Lijun Ye, Yueping Xi, Chang Chen, Feng Xiong, Hongzhan Zhang, Xuejin Wang

## Abstract

Non-obstructive azoospermia (NOA) represents the most severe form of male infertility, severely limiting a patient’s prospects for biological fatherhood when surgical retrieval fails. However, the true biological limits of NOA remain obscured by the inherent limitations of conventional gamete recovery protocols: standard centrifugation frequently causes substantial cell loss, masking extremely rare sperm, while surgical interventions are constrained by spatial sampling biases. Here we report SpermSeek, an integrated AI-guided microfluidic platform for real-time, non-destructive isolation of single sperm directly from semen. Operating at scalable throughput (0.36 mL/h), the system achieves 98.3% detection precision and a 95.5% target encapsulation efficiency, suppressing background debris. In a 59-patient NOA cohort, SpermSeek detected morphologically identifiable sperm in 64.4% (38/59) of cases, spanning diverse genetic etiologies, including AZFb/c microdeletions, and severe histopathological phenotypes, such as Sertoli-cell-only syndrome (SCOS). Notably, among a sub-cohort of 41 patients who remained consistently sperm-negative despite prior medical or micro-TESE interventions, our platform identified gametes in 53.7% (22/41) of these cases. Comprehensive safety profiling in healthy human donors and wild-type mice confirmed that processed sperm retain high DNA integrity and epigenomic concordance (r=0.98), supporting transgenerational developmental stability in mice. Furthermore, in a 26-patient validation cohort, SpermSeek recovered rare sperm in 11 cases. Utilizing gametes from a subset (n=5), we demonstrated their capacity to support early human embryogenesis, yielding high-quality cleavage-stage embryos with confirmed genomic euploidy. This work establishes a highly sensitive framework for re-examining the biological limits of human spermatogenesis, laying the foundation to expand autologous reproductive options for patients refractory to conventional retrieval protocols.

## Introduction

Male factor infertility affects approximately 50% of infertile couples worldwide, with non-obstructive azoospermia (NOA) representing its most severe form[1-3]. Characterized by the absence of sperm in ejaculated semen due to intrinsic spermatogenic failure, NOA encompasses diverse etiologies (such as Y-chromosome microdeletions[4]) and complex histopathological patterns including Sertoli-cell-only syndrome (SCOS)[5], tubular hyalinization[6], maturation arrest[7] and hypospermatogenesis[8]. For these patients, the failure to recover sperm following standardized endocrine therapies or microdissection testicular sperm extraction (micro-TESE) conventionally severely limits their autologous reproductive options[2, 9-11].

This conventional clinical consensus, however, is heavily dictated by the inherent limitations of standard World Health Organization (WHO) protocols[12]. Although the manual examination of centrifuged seminal pellets serves as the first-line screening method, it lacks sufficient sensitivity to reliably recover extremely scarce sperm due to an unavoidable physical paradox. At low speeds, rare gametes fail to sediment and remain suspended in viscous supernatants, making manual microscopic detection physically intractable despite repeated enrichment cycles[13, 14]. Conversely, the higher centrifugation speeds necessary to force sedimentation inflict mechanical stress and significant processing-induced cell loss[15]. Similarly, although micro-TESE represents the surgical gold standard for non-obstructive azoospermia (NOA), its efficacy remains fundamentally constrained by spatial sampling limitations; localized tissue excisions frequently miss highly heterogeneous, focal islands of preserved spermatogenesis within the testes.

Advanced high-resolution cell sorting technologies—including flow cytometry[16], magnetic capture[17], and conventional microfluidics[18-20] —are widely established in broader biomedical fields. However, these systems are primarily engineered to enrich abundant subpopulations, which makes them inherently unsuitable for clinical gamete recovery. Specifically, flow cytometry and magnetic capture typically entail exogenous labeling, intense laser interrogation, or high-shear column passages. These requirements introduce biological safety concerns[21, 22] and substantial processing-induced cell loss[23], which largely restrict their clinical application for rare gamete recovery. In contrast, while conventional microfluidic techniques offer label-free alternatives, they predominantly depend on active sperm motility for sorting—a functional trait that is frequently compromised or entirely absent in NOA samples[18, 19]. Furthermore, conventional continuous-flow microfluidic systems often dilute the recovered sperm into large output volumes, complicating subsequent single-cell retrieval and downstream manipulation[18, 24]. Consequently, a critical technological gap remains: a framework capable of continuously screening large semen volumes, identifying individual sperm at extreme rarity, and recovering them in a controlled, biologically preserved manner.

A recent proof-of-concept study[25] established the operational feasibility of AI-assisted microfluidics for single-sperm identification and retrieval. However, while providing an essential first step, this application focused on a patient with a documented history of successful gamete detection—having previously yielded rare sperm through both extensive manual screening and multiple testicular sperm extraction (TESE) surgeries. Thus, an important question lies at the intersection of reproductive biology and clinical technology: does a persistent ’zero-sperm’ diagnosis following repeated medical and surgical interventions, such as micro-TESE, reflect a true biological depletion of germ cells? Or do functionally and genomically intact gametes persist in a subset of these patients, merely obscured by the intrinsic detection limits of current clinical gold standards?

Here, we resolve this ambiguity by profiling a 59-patient historically sperm-negative NOA cohort using SpermSeek—an integrated AI-guided microfluidic platform for non-destructive rare-sperm recovery. We reveal that the ’zero-sperm’ diagnosis is frequently a technological artifact of centrifugation loss and spatial sampling biases, rather than an absolute biological dead-end. Crucially, we demonstrate that intact gametes persist even in patients unresponsive to extensive medical or surgical interventions, and confirm that these safely recovered gametes retain the capacity to generate genomically euploid cleavage-stage embryos. Ultimately, this work establishes a highly sensitive framework for re-examining the biological limits of human spermatogenesis, laying the foundation to expand autologous reproductive options for patients refractory to conventional retrieval protocols.

## Results

### 1. SpermSeek: A closed-loop, vision-activated microfluidic platform for non-destructive rare-sperm isolation

To overcome the physical limitations of conventional rare-cell recovery, we engineered SpermSeek, an integrated microfluidic platform designed for the automated, non-destructive isolation of target sperm directly from complex NOA semen (Fig. 1). The operational workflow is initiated by high-throughput dynamic optical imaging and deep learning-based recognition (Fig. 1a), which trigger ultra-low-latency microfluidic actuation to isolate single sperm under continuous flow (Fig. 1b). Building upon this core hardware–software architecture, we established a translational pipeline. This encompasses the systematic morphological profiling of clinical NOA cohorts (Fig. 1c), followed by multi-omic and transgenerational safety evaluations of the microfluidic sorting mechanism using healthy human donors and wild-type mice (Fig. 1d). Finally, the pipeline concludes with the functional validation of recovered rare NOA sperm through standard embryogenic assessments (Fig. 1e).

**Figure 1.**
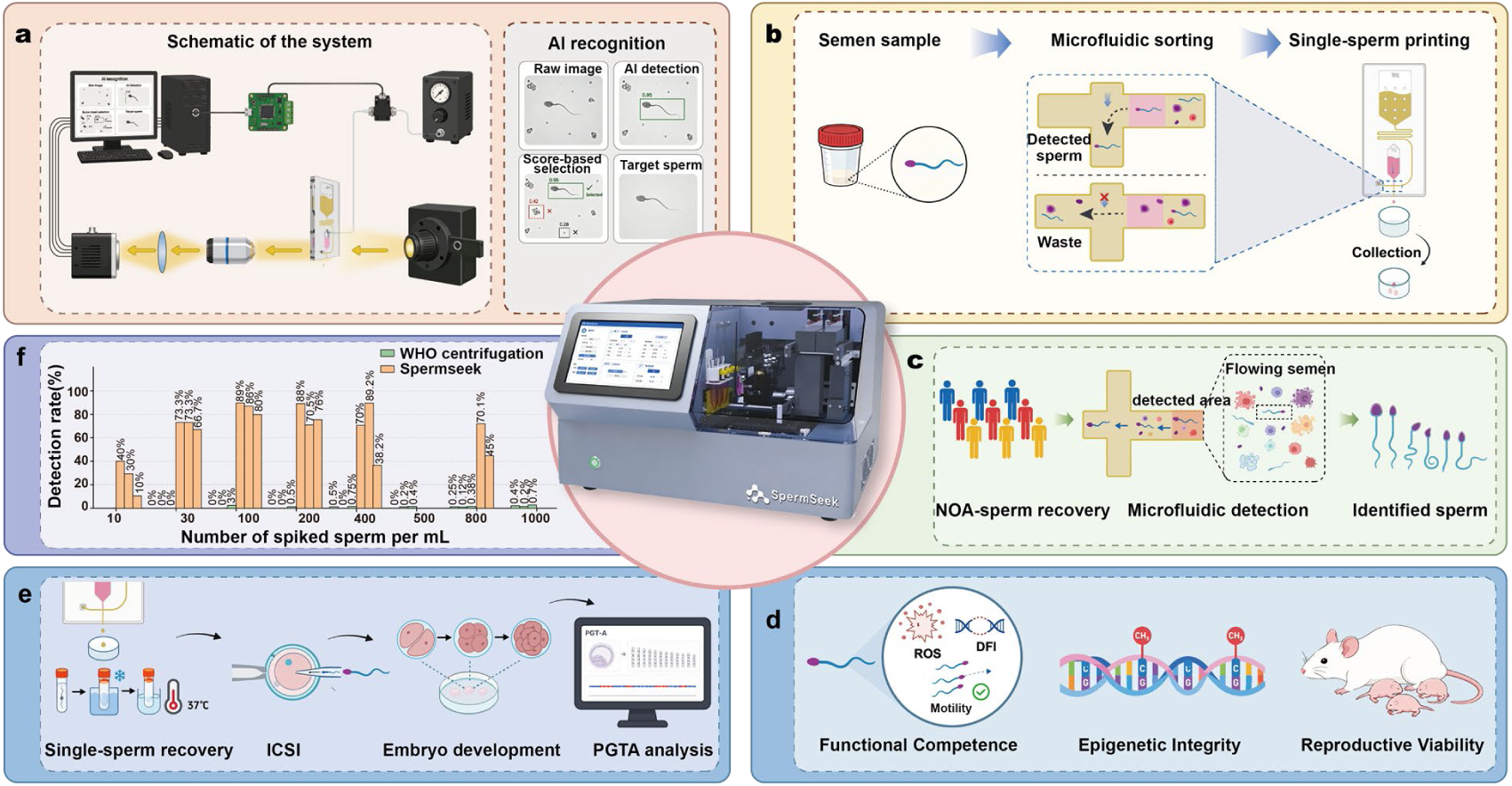
The SpermSeek platform and NOA clinical translation pipeline. (Center) 3D rendering of the fully integrated SpermSeek instrument. (a) System architecture integrating core microfluidic hardware and deep learning for real-time target recognition. (b) Closed-loop microfluidic actuation for single-sperm droplet printing. (c) Clinical screening workflow for a 59-patient NOA cohort. (d) Functional, genetic, and epigenetic safety validation pipeline using healthy human and wild-type mouse sperm. (e) Downstream clinical recovery and fertilization validation of rare NOA sperm. (f) End-to-end detection benchmarking via spiking assays.

At the hardware level, SpermSeek operates under a clinically scalable, continuous-flow regime (volumetric throughput up to 0.36 mL/h). The system integrates a software-defined, GPU–CPU heterogeneous computing architecture that enables low-latency, inference-driven closed-loop control for targeted sperm encapsulation via a microfluidic nozzle. This ultra-fast actuation eliminates motion-induced spatial mismatches during high-speed flow (approximately 21 mm s⁻¹), ensuring that individual sperm are extracted with minimal mechanical perturbation.

Concurrently, systematic evaluation of the algorithmic layer demonstrated that the AI-driven vision module maintains high precision even in the presence of dense background noise typical of NOA samples (e.g., spermatogenic cells, leukocytes, and tissue debris). Powered by a lightweight YOLOv8n object detection architecture optimized for real-time inference, the integrated model was evaluated on an independent test set. It achieved a baseline mean Average Precision (mAP@0.5) of 86.7%, a precision of 79.1%, and a recall of 77.6%. Crucially, rather than relying solely on static algorithmic benchmarking, we evaluated the vision module using task-driven clinical sorting criteria. In dynamic microfluidic workflows, rigid bounding-box constraints are superseded by the requirement for inclusive droplet encapsulation, where capturing morphologically representative ’sperm-like’ entities guarantees the retrieval of actionable gametes. Evaluated under these practical biological parameters, the model achieved a precision of 98.3% and a recall of 96.4%. This corresponds to a false-negative rate of 3.6% and a low false discovery rate of 1.7%. Ultimately, this discrimination isolates high-purity sperm by suppressing false triggers from background debris.

To benchmark this end-to-end recovery efficacy against the severe cell loss inherent to standard clinical protocols (e.g., WHO-recommended extended centrifugation), we designed deterministic spiking experiments. These bypassed stochastic dilution errors by introducing exact sperm gradients into sperm-free seminal plasma (prepared via high-speed centrifugation of healthy donor samples), establishing an absolute ground truth. The results exposed a processing-induced attrition inherent to standard WHO centrifugation: across spiked gradients ranging from 30 to 1,000 sperm, manual microscopic examination of the resulting pellets yielded near-zero recovery. Even at the spike of 1,000 sperm, a maximum of merely 7 cells were recovered across triplicate experiments (a <0.7% recovery rate), while lower-concentration cohorts repeatedly dropped to absolute zero (Fig. 1f).

In contrast, by synergizing continuous-flow microfluidic screening with high-precision AI target recognition, SpermSeek addresses the physical limitations of rare sperm detection. Across the discrete tested gradients between 30 and 800 sperm per mL, the system maintained an end-to-end recovery rate averaging between 57.5% and 85%. Furthermore, even at the extreme physical threshold of 10 sperm per mL, the platform still achieved an average recovery of 26.7% (Fig. 1f). By overcoming the irreversible gamete loss inherent to conventional enrichment protocols, the system operates in a regime that is intrinsically unrecoverable by standard means. When applied to unspiked, extreme NOA clinical samples, this continuous-flow pipeline demonstrates the capacity to retrieve actionable gametes—isolating down to a single viable sperm hidden within a 2 mL volume of patient semen.

### 2. Microfluidic design for precise and stable single-sperm droplet printing

To achieve rapid and non-destructive single-sperm printing, we engineered a microfluidic platform built upon an orthogonal flow-decoupling strategy (Fig. 2a). The system utilizes a primary horizontal channel for the continuous, low-shear screening of the raw semen sample. This screening flow is orthogonally intersected by a vertical printing axis at a localized recognition junction. Under continuous operation, seminal plasma and unrecognized debris flow toward the waste outlet. However, upon real-time algorithmic identification of the sperm, an ultra-fast, tunable pressure pulse (typically 2–4 ms in duration) drives the vertical printing buffer to orthogonally redirect the targeted cell out of the primary streamline in <0.33 ms. Immediately following this redirection, the sperm is encapsulated and ejected within a nanoliter droplet via an integrated micro-nozzle (Fig. 2b). By decoupling the continuous macroscopic sample screening from the discrete microscopic droplet ejection, this cross-junction architecture minimizes hydrodynamic disturbance, preserving sperm structural integrity.

**Figure 2.**
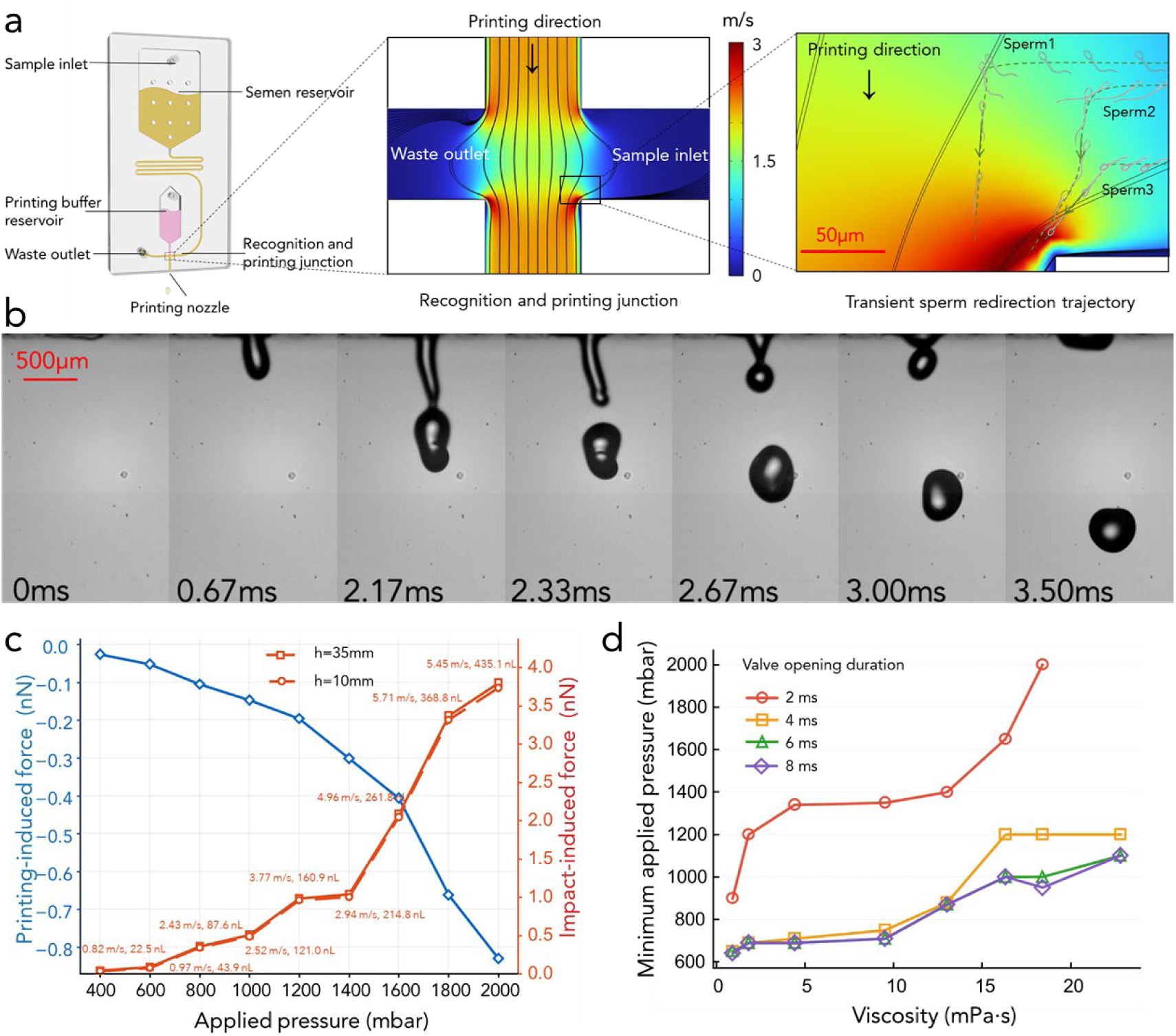
Mechanistic basis of non-destructive single-sperm printing. (a) 3D schematic rendering of the microfluidic chip. Magnified views of the cross-junction feature computationally resolved streamlines delineating the background flow field. Against this simulated fluidic environment, superimposed experimental frames trace the transient (<0.33 ms) orthogonal redirection of a single sperm from horizontal continuous flow to vertical droplet ejection. (b) High-speed micrographs capturing the single-droplet (40 nL) generation and jetting dynamics. (c) Simulated peak hydrodynamic and impact forces acting on a spherical sperm surrogate during on-chip flow redirection and off-chip droplet deposition. (d) Operational phase diagram mapping the minimum actuation pressures required for stable droplet ejection across varying fluid viscosities (evaluated via viscosity-matched glycerol-water surrogates) and pulse durations.

To evaluate the mechanical stress exerted on sperm during the printing process, we developed a multiphysics computational model—parameterized by our experimental conditions—to reconstruct the transient hydrodynamic microenvironments of both the on-chip orthogonal flow redirection and the off-chip droplet impact against the substrate. As shown in Fig. 2c, during the initial on-chip flow redirection, increasing the printing pressure from 40 kPa (corresponding to a measured jet-tip velocity of 0.82 m s⁻¹) to 200 kPa (5.45 m s⁻¹) amplified the peak printing-induced force acting on the spherical sperm surrogate from approximately 0.026 nN to 0.83 nN.

Concurrently, across practical dispensing heights (10–35 mm), the peak impact-induced force during droplet impact onto a rigid substrate was estimated to range from 0.03 to 3.80 nN. High-speed imaging confirmed that this on-chip flow-redirection event is transient, restricting the duration of peak mechanical exposure to a sub-millisecond temporal window.

To provide a clinical baseline, a spherical sperm surrogate (diameter 5 μm, density 1050 kg m⁻³) subjected to WHO-recommended 3000×*g* centrifugation endures a sustained individual inertial load of approximately 2 nN for 15 minutes[12]—a baseline stress that is compounded by mechanical shear forces, physical compaction within the pellet and the consequent elevation of reactive oxygen species (ROS) during pellet compaction at the tube bottom[15, 24, 26]. Although our maximum impact-induced forces (up to 3.80 nN) momentarily exceed this 2 nN static baseline, this exposure is sub-millisecond, thereby generating a negligible mechanical impulse. Furthermore, the peak printing-induced forces during the on-chip flow redirection (<0.83 nN) remain below this baseline. SpermSeek reduces the mechanical exposure duration from 15 minutes to <0.33 ms and eliminates pellet-induced compressive damage through its continuous-flow regime. This minimization of cumulative mechanical stress provides the physical basis for preserving the structural and functional integrity of fragile NOA sperm populations.

Beyond intrinsic mechanical safety, the platform demonstrates operational stability across the broad rheological heterogeneity of clinical samples. Profiling 23 clinical semen specimens revealed a wide viscosity span ranging from 1.07 to 20.1 mPa·s. To decouple the effects of bulk viscosity from stochastic perturbations caused by seminal debris, we mapped the system’s operational phase boundary using viscosity-matched glycerol-water surrogates. By evaluating the critical actuation pressures required for stable droplet ejection across a 2–8 ms pulse-duration regime (Fig. 2d), we confirmed that the platform maintains consistent, occlusion-free droplet generation. For practical pulse durations (≥4 ms), the minimum applied pressure remained tightly bounded below 1200 mbar across the entire clinical viscosity spectrum, ensuring reliable operation without the need for extreme system pressurization. Capitalizing on this wide operational window, we translated these physical limits into a discrete set of standardized actuation profiles, eliminating the need for iterative, sample-specific pressure calibration during all subsequent clinical processing.

### 3. Systematic profiling of rare sperm across the diverse spectrum of NOA

To systematically map the clinical boundaries of rare sperm occurrence, we screened a 59-patient NOA cohort—53 of whom were historically sperm-negative—using the SpermSeek platform. Overall, the system identified morphologically intact gametes in 64.4% (38/59) of the cases, with concentrations ranging from 1 to 49 sperm mL⁻¹ (median: 3 sperm mL⁻¹; mean: ∼8 sperm mL⁻¹; Fig. 3a).

**Figure 3.**
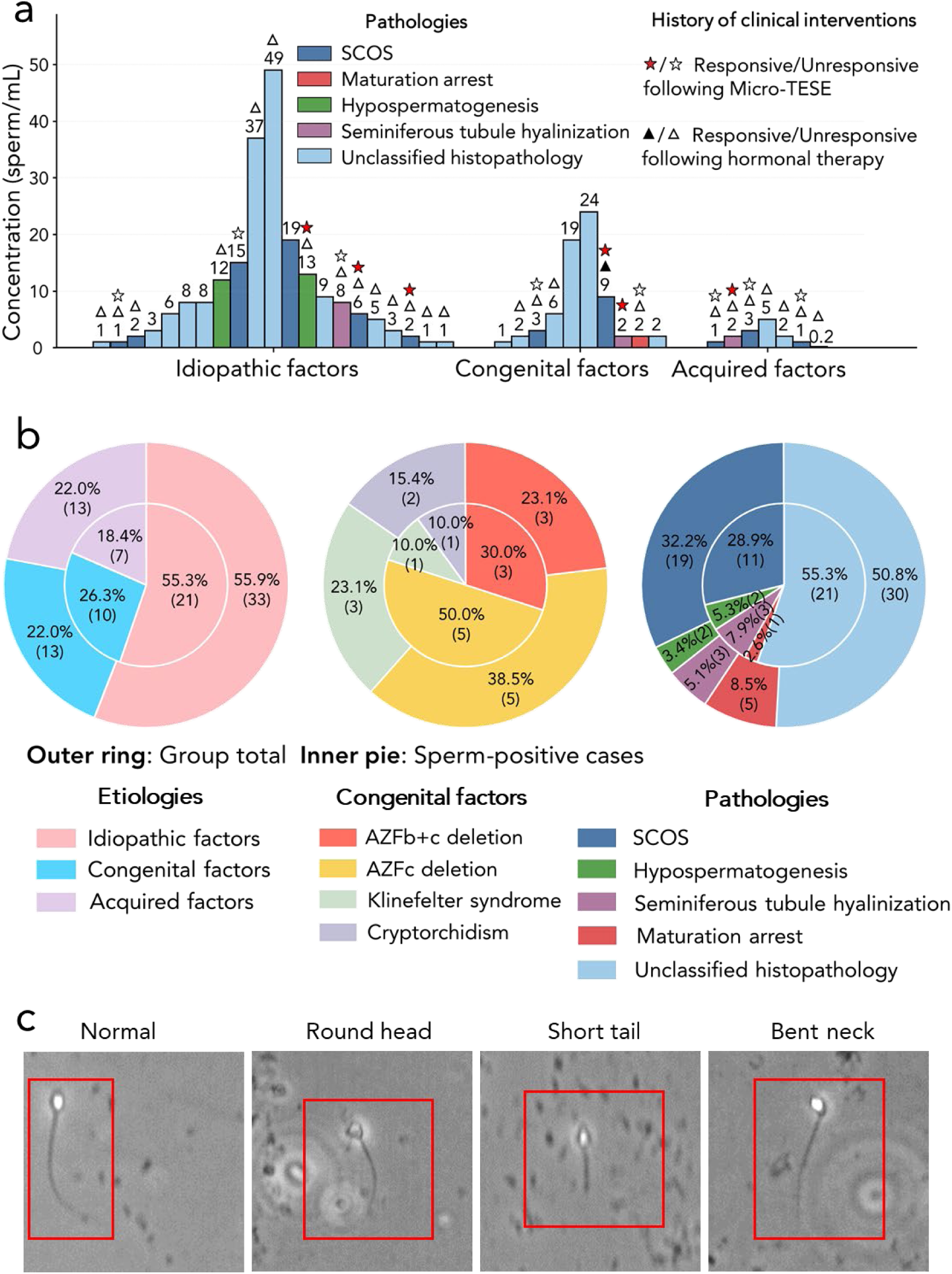
Systematic profiling of rare sperm in a clinical azoospermic cohort. (a) Patient-specific rare sperm concentrations stratified by etiological factors and underlying pathologies. Symbols denote the patient’s history and responsiveness to prior clinical interventions (micro-TESE and hormonal therapy). (b) Distribution landscapes of etiologies (left), congenital genetic factors (middle), and histopathologies (right). Outer rings denote the total patient cohort (N = 59); inner pies represent the sperm-positive subgroup (n = 38). (c) Representative micrographs of retrieved sperm exhibiting structurally normal and anomalous morphologies (e.g., round head, short tail, bent neck).

A substantial portion of our cohort represents a previously treated demographic (Fig. 3a). Specifically, a subset of 47 patients had previously undergone targeted interventions. Within this treated group, 87.2% (n = 41) received pharmacological therapy, 51.1% (n = 24) underwent micro-TESE, and 38.3% (n = 18) received a combination of both. To investigate whether ultra-low-abundance spermatogenesis persists within this consistently unresponsive demographic, we stratified these treated patients based on their historical recovery outcomes. Among the 41 patients who remained consistently sperm-negative despite these interventions, the SpermSeek platform identified gametes in 22 cases (53.7%; Fig. 3a, symbols). The remaining 6 treated patients, who had previously yielded rare sperm via surgical extraction (micro-TESE, with one patient also responding to hormonal therapy), served as clinical positive controls. Historical records for these patients documented marginal gamete yields, ranging from a single immotile spermatozoon in the entire surgical specimen to a maximum of 10 sperm (4 motile, 6 immotile) per 20 high-power fields. Under our rigorous morphological verification protocol, SpermSeek demonstrated 100% concordance in this subgroup (6/6), identifying target concentrations of 2–13 sperm mL⁻¹ from the ejaculate.

We next profiled the clinical features of the cohort (N = 59) to assess the occurrence of identifiable sperm across specific etiologies, congenital genetic factors, and histopathological parameters (Fig. 3b). Etiologically, the cohort comprised idiopathic azoospermia (55.9%), congenital factors (22.0%), and acquired conditions (22.0%). Across these distinct clinical origins, rare gametes were found to persist at comparable frequencies, with positive detections in 63.6% (21/33) of idiopathic, 76.9% (10/13) of congenital, and 53.8% (7/13) of acquired cases.

Further characterization of the congenital genetic subgroup (Fig. 3b, middle) detected scarce sperm in Y-microdeletion carriers. Prior studies reported very low sperm retrieval rates with routine semen centrifugation or micro-TESE[27]. We identified sperm in all three patients with complete compound AZFb+c deletions, a genotype rarely yielding gametes via standard methods. The AZFc-related group contained cases with partial AZFb plus AZFc deletions and isolated AZFc deletions. All five patients in this subgroup were azoospermic on routine testing, yet low-concentration sperm were detected by SpermSeek. Sperm concentrations across all genetic subgroups ranged from 1 to 24 sperm mL⁻¹.

At the histopathological level (Fig. 3b, right), patients exhibited significant testicular impairment, primarily including SCOS (32.2%), maturation arrest (8.5%), seminiferous tubule hyalinization (5.1%) and hypospermatogenesis (3.4%). Trace-level spermatogenesis remained evident even within the most severe phenotypes. In SCOS—where "complete" pathology theoretically implies an absence of germline cells[28]—morphologically intact sperm (1–19 sperm mL⁻¹) were identified in 11 of 19 diagnosed patients. Similarly, gametes (2–8 sperm mL⁻¹) were identified in all three patients diagnosed with tubular hyalinization, a condition characterized by severe tubular degeneration[6].

Morphologically, the detected cells exhibited substantial heterogeneity (Fig. 3c), comprising both sperm with preserved head–tail architecture and structurally abnormal forms (e.g., irregular heads, coiled tails, or abnormal neck–tail junctions). To define the biological boundaries of true gamete occurrence, we enforced strict, expert-verified morphological criteria. In the remaining 35.6% (21/59) of patients, no cells met these rigorous criteria. Although rare "sperm-like" structures lacking complete architectural integrity were occasionally observed, they were conservatively excluded from all quantitative and functional analyses to ensure rigorous diagnostic stringency.

Collectively, these findings reveal that rare populations of sperm persist across diverse NOA pathologies and major etiologies at a higher prevalence than traditionally assumed. The ability to uncover these elusive gametes is largely attributed to SpermSeek’s volumetric screening of the sample (typically 1–3 mL). When processing highly heterogeneous NOA samples requiring brief pre-centrifugation, we frequently recovered rare sperm directly from the supernatant. This observation highlights a critical physical constraint in rare-cell retrieval: because manual microscopic screening is physically incapable of processing large-volume liquids, conventional protocols are forced to rely on prolonged, high-speed centrifugation (such as the WHO 15-minute standard) to concentrate cells into a pellet, inherently risking mechanical damage. In contrast, SpermSeek’s scalable throughput allows for the continuous screening of the entire buoyant supernatant following a brief pre-centrifugation. This bypasses the physical paradox, recovering elusive gametes while circumventing the need for pelleting forces.

### 4. Functional, epigenetic, and *in vivo* reproductive validation of SpermSeek-processed sperm

Before clinical application, it is imperative to verify that the SpermSeek isolation process does not induce secondary mechanical or oxidative damage. First, we evaluated the basal functional parameters and cellular integrity of SpermSeek-processed sperm from healthy human donors. Quantitative paired analyses revealed that key physiological metrics—including total motility (TM), progressive motility (PM), reactive oxygen species (ROS) levels, and DNA fragmentation index (DFI)—remained unaltered compared to unmanipulated controls (Fig. 4a, all *P* > 0.05). These results confirm that the microfluidic processing preserves gamete viability without measurably compromising cellular quality.

**Figure 4.**
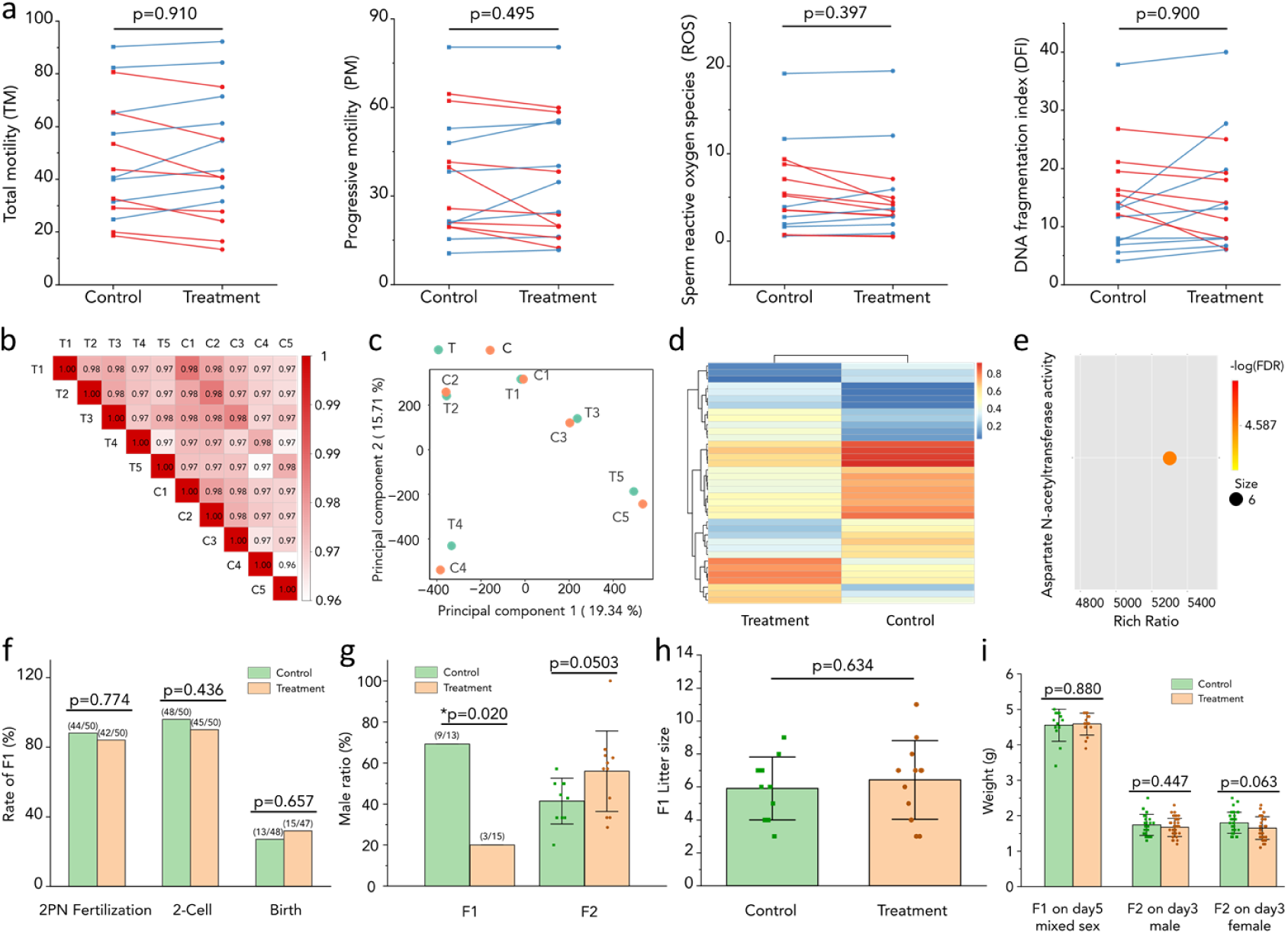
SpermSeek preserves the functional, epigenetic, and reproductive integrity of sperm. All analyses compare SpermSeek-processed with unmanipulated control sperm. (a) Paired analysis of sperm quality parameters (Total motility, progressive motility, ROS, and DNA fragmentation index) in healthy donors (n = 16). Red/blue lines denote individual decreases/increases. (b–e) DNA methylome profiling of sperm from healthy donors (n = 5 paired replicates). (b) Pearson correlation and (c) principal component analysis (PCA) demonstrating high global epigenomic concordance and clustering by individual origin rather than processing status. (d) DMR heatmap and (e) Gene Ontology enrichment of hypermethylated DMRs. (f–i) Transgenerational safety in a wild-type mouse ICSI model. (f) Developmental milestones (F1 progeny derived from n = 2 surrogate mothers per group). (g) Male sex ratios (F1, F2). (h) F1 litter sizes. (i) Postnatal weights of F1 (mixed-sex) and F2 (sex-stratified) offspring.

Having confirmed the preservation of physical and functional traits, we next investigated whether the microfluidic isolation induced any latent molecular alterations. To this end, we profiled the high-resolution epigenetic landscape of human sperm via whole-genome bisulfite sequencing (WGBS; n = 5 paired replicates). Global DNA methylome profiles exhibited high concordance across all samples (r≥ 0.96, paired mean r = 0.98; Fig. 4b). Principal component analysis (PCA; PC1 19.34%, PC2 15.71%) revealed that samples clustered by individual donor origin rather than by processing status (Fig. 4c), demonstrating that microfluidic processing introduces less epigenomic variance than baseline inter-individual heterogeneity. Using a stringent analysis pipeline, we identified only 37 differentially methylated regions (DMRs) between processed and control sperm (Fig. 4d). Gene Ontology (GO) molecular function enrichment of the hypermethylated DMRs identified a single association with aspartate N-acetyltransferase activity (Fig. 4e), while hypomethylated regions mapped to tRNA-dimethylallyltransferase and D-ribulokinase activities. None of these specific metabolic functions are linked to spermatogenesis, genomic imprinting, or embryonic development.

With both phenotypic and epigenomic stability established *in vitro*, we validated the *in vivo* reproductive outcomes and transgenerational safety of SpermSeek-processed gametes via ICSI in a wild-type murine model. Both 2PN fertilization and cleavage rates were comparable between the control and processed sperm groups (2PN fertilization rate: 88.0% vs. 84.0%, *P* = 0.774; 2-Cell rate: 96.0% vs. 90.0%, *P* = 0.436, Fig. 4f). Following embryo transfer, live birth rates showed no significant difference (27.1% vs. 31.9%, *P* = 0.657).

While we observed a statistical shift in the F1 male sex ratio (Fig. 4g, *P* = 0.02), this divergence was driven by an unusually high baseline in the control group (69.2%) combined with a lower ratio in the processed group. This fluctuation is consistent with natural litter-to-litter biological variance rather than a systematic, technology-induced sex selection bias. This conclusion is corroborated by the F2 generation, where continuous transgenerational mating fully normalized the male ratio to expected comparable levels (Fig. 4g).

Furthermore, transgenerational reproductive capacity was preserved. The mating of F1 animals yielded a total of 59 and 77 F2 pups in the control and processed cohorts, respectively, and their average litter sizes were comparable between groups (Fig. 4h). Despite the F1 sex ratio variance, systemic development of the F1 offspring remained stable: birth weights showed no significant difference compared to controls (mean ± SD: 4.55 ± 0.45 g vs. 4.59 ± 0.31 g, *P* = 0.880), and their sex-specific postnatal growth trajectories were comparable to those of the respective control groups. Furthermore, this transgenerational stability extended to the F2 cohort, where sex-stratified postnatal weights also remained indistinguishable from controls (Fig. 4i).

Collectively, multi-scale evidence—spanning human cellular physiology (Fig. 4a), high-resolution epigenomics (Fig. 4b–e), and transgenerational mammalian development (Fig. 4f–i)—demonstrates that the SpermSeek platform preserves reproductive potential without compromising genomic or developmental stability.

### 5. Embryogenic potential and genomic stability of rare sperm in NOA

Having established the functional, epigenomic, and transgenerational safety of the platform, we sought to address a fundamental biological question: whether rare sperm hidden in the ejaculate of otherwise ’sperm-negative’ NOA patients retain the intrinsic capacity to initiate and support early embryogenesis. For cases traditionally presumed to lack clinically actionable spermatogenesis, the developmental and genomic integrity of their hidden gametes remains unexamined, as their high-purity recovery was previously unachievable. To functionally evaluate the embryogenic potential of these elusive gametes, we applied our integrated platform to an *in vitro* validation cohort of 26 NOA patients, comprising cases previously identified as sperm-positive via our detection module (Section 3) alongside newly recruited NOA patients undergoing their first SpermSeek evaluation.

During the physical recovery phase, we deliberately tracked and isolated only the most morphologically intact sperm via on-chip continuous flow to maximize subsequent ICSI viability (Fig. 5a). Applying this stringent criterion across the validation cohort of 26 individual patients, sperm were recovered in 11 cases (a patient-level recovery efficiency of 42.3%; Fig. 5b). The divergence between the 64.4% diagnostic detection rate (Section 3) and this experimental recovery rate is primarily driven by stochastic sampling limitations—arising from processing only 1-mL aliquots of ultra-low concentration samples—as well as the stricter morphological threshold compared to basic diagnostic requirements.

**Figure 5.**
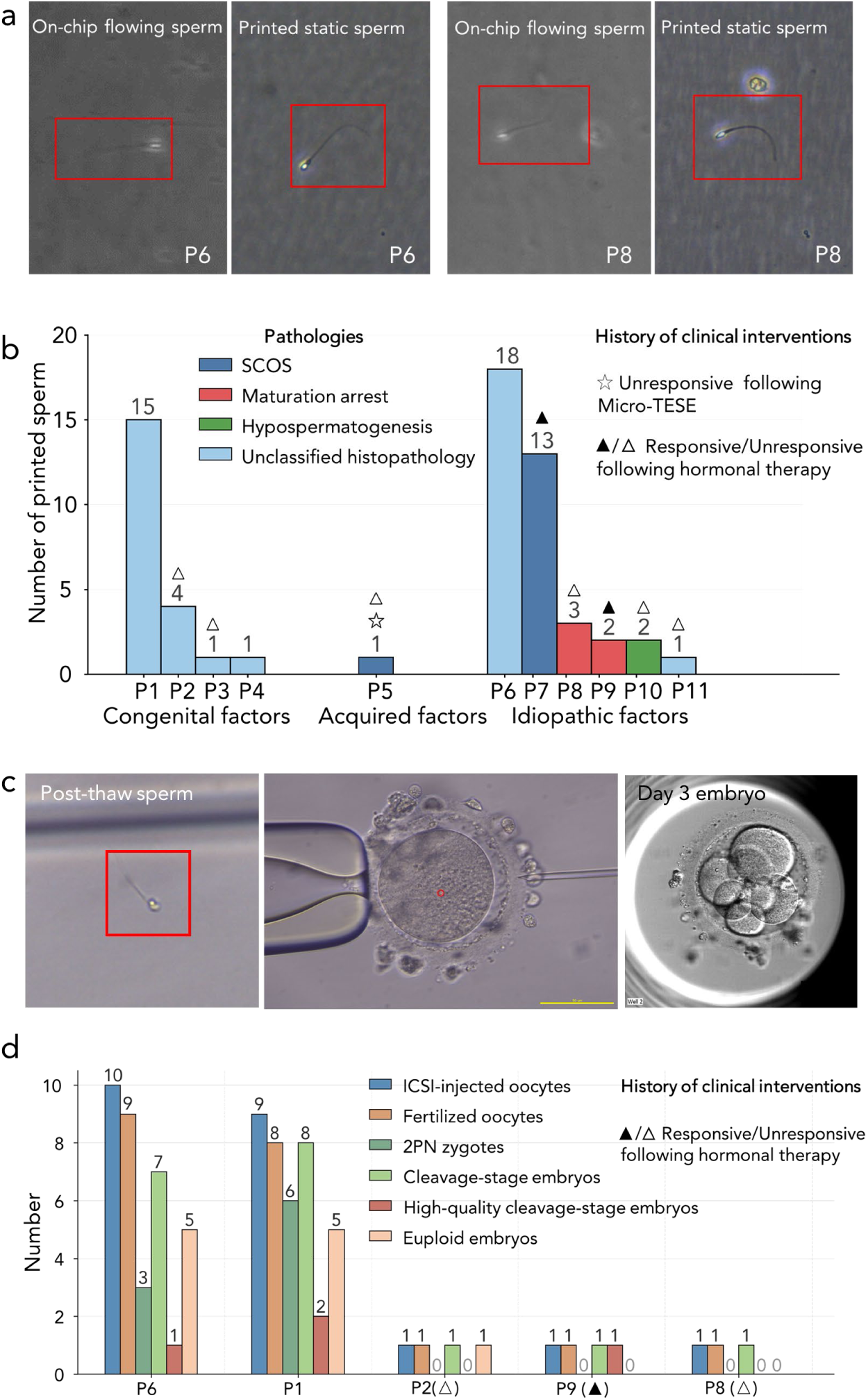
Clinical recovery and embryogenic potential of rare NOA sperm. (a) Morphological tracking of individual sperm (patients P6, P8) during on-chip continuous flow and post-isolation. (b) Sperm recovery yields across individual patients (P1–P11), stratified by underlying etiologies and pathologies. (c) Clinical workflow showing a post-thaw recovered sperm, the ICSI procedure, and a resulting day-3 embryo. (d) Post-ICSI developmental outcomes, including fertilization metrics and euploid embryo yields validated via whole-embryo preimplantation genetic testing for aneuploidy (PGT-A).

Recovered sperm spanned diverse clinical etiologies at ultra-low concentrations, including patients with complete compound AZFb+c deletions (0.5 sperm mL^-1^), partial AZFb plus AZFc deletions, and isolated AZFc deletions (4–34 sperm mL⁻¹), as well as those with prior testicular trauma (0.5 sperm mL^-1^), cryptorchidism (1 sperm mL^-1^), and idiopathic azoospermia (1–36 sperm mL^-1^). Recoveries included individuals who had remained consistently sperm-negative despite prior interventions, such as endocrine therapy alone or in combination with micro-TESE (indicated by symbols, Fig. 5b). These findings indicate that non-invasive sperm recovery is feasible even in the severe NOA phenotypes traditionally presumed to preclude spermatogenesis.

To determine whether the recovered rare gametes maintain structural stability for downstream clinical applications, sperm from 5 patients were cryopreserved and thawed. Post-thaw microscopic evaluation confirmed that the isolated gametes maintained structural and morphological integrity (Fig. 5c). To comply with ethical protocols, ICSI was restricted to *in vitro* research using anonymized healthy donor oocytes, and the resulting embryos were prohibited from clinical transfer. Across 22 injected oocytes, the overall fertilization rate was 90.9% (20/22), yielding 9 normal two-pronuclear (2PN) zygotes (Fig. 5d) and 4 high-quality cleavage-stage embryos (Fig. 5c, d).

Regarding extended *in vitro* culture, the high-quality cleavage embryo rate was 18.2% (4/22) per injected oocyte (Fig. 5d). However, continuous observation from Day 3 to Day 7 did not result in progression to the blastocyst stage. The clinical context and potential factors contributing to this *in vitro* post-cleavage outcome—particularly the constraints of utilizing ultra-low quantities of unselected gametes—are further evaluated in the Discussion.

To evaluate genomic stability—a critical unknown for gametes recovered from NOA —all 18 developing embryos (comprising the 4 high-quality cleavage-stage embryos and 14 lower-grade embryos with delayed or atypical cleavage) were subjected to whole-embryo lysis for PGT-A profiling. The remaining 4 injected oocytes were excluded due to failed fertilization or unsuccessful cleavage. 61.1% (11/18) of the profiled embryos were confirmed as euploid with normal copy number profiles (Fig. 5d). The remaining 7 embryos exhibited mosaicism or aneuploidy.

Together, these findings provide functional evidence that rare sperm recovered from NOA patients—including subtypes traditionally considered incapable of yielding recoverable gametes—retain the intrinsic capacity to support fertilization and early embryonic development. Genomic euploidy in over half of the resulting embryos supports the biological viability of these previously inaccessible gametes, and validates the capability of our platform for high-purity, single-cell functional recovery.

## Discussion

While surgical extraction and pharmacological therapy have salvaged fertility for a subset of NOA patients, those unresponsive to these measures are presumed to lack reproductive potential. Here, we demonstrate that in many of these unresponsive cases, the presumed absence of sperm is a consequence of conventional detection limits rather than a complete loss of functional spermatogenesis. By integrating deep learning with ultra-low-latency microfluidic actuation, the SpermSeek platform breaches the physical "blind spot" of conventional diagnostics. Our findings reveal that rare gametes persist in a substantial proportion of NOA patients, and such gametes can retain early embryogenic competence. Together, these findings challenge the established diagnostic boundaries of presumed complete spermatogenic failure.

Translating this micro-scale detection into reliable clinical utility requires a fundamental transition in engineering execution. Unlike highly customized laboratory prototypes that demand complex manual calibration, SpermSeek functions as a scalable, operator-independent integrated system. Within this integrated architecture, the current volumetric throughput (6 µL min⁻¹, requiring approximately 2.8 h per 1 mL ejaculate) represents a biophysical trade-off to maximize clinical fidelity. Although our platform could achieve higher flow rates (e.g., 8 µL min⁻¹) by utilizing brightfield imaging (with exposure times of 15 µs at 260 fps), this optical compromise degrades the AI-recognition signal-to-noise ratio in dense seminal debris, and the accelerated flow risks unacceptable rare-target loss. To maximize algorithmic specificity, our platform utilizes phase-contrast microscopy of native, unwashed semen. This dictates longer exposure times (300–500 µs) and caps the operational frame rate at 180 fps. In the context of NOA—where retrieving even a single viable gamete dictates clinical actionability—maximal diagnostic sensitivity and damage-free preservation must supersede raw processing speed. Future iterations integrating high-intensity pulsed LED illumination will reduce exposure times, enabling higher operational frame rates and elevated volumetric throughput while preserving critical optical contrast.

Beyond technological utility, our systematic screening of this historically sperm-negative clinical cohort offers essential biological insights into male germline persistence. Our consistent detection of sperm across a broad spectrum of NOA subtypes—including AZFb/c microdeletions, Sertoli cell-only syndrome (SCOS), and tubular hyalinization—refines the prevailing clinical assumption that such severe histological phenotypes or pathogenic genetic aberrations prevent gametogenesis.

Our findings demonstrate that these previously inaccessible gametes are not merely structural remnants, but can retain essential functional competence for embryogenesis. Following clinical cryopreservation and thawing, utilizing these rare gametes for human ICSI yielded a high fertilization rate (90.9%) and supported early embryogenesis, producing high-quality Day 3 cleavage-stage embryos, and confirming genomic euploidy in a substantial proportion of the cohort. Particularly for NOA patients consistently diagnosed as ’sperm-negative’ under standard clinical evaluations, the generation of chromosomally normal cleavage embryos provides a critical proof-of-concept for clinical translation—shifting their prognosis from a presumed complete loss of functional spermatogenesis to the realm of actionable reproductive potential.

While extended *in vitro* monitoring did not yield blastocyst formation by Day 7, this outcome is partially attributable to the extreme gamete scarcity in NOA, which restricts the rigorous morphological and kinetic selection typically employed in standard ICSI[29]. Furthermore, our exclusive research-based reliance on a double-cryopreserved model (frozen NOA sperm and frozen donor oocytes) is known to exert additional stress on developmental potential[30]. However, the absence of *in vitro* blastocysts does not negate the immediate clinical utility of these gametes. In established ART practice for poor-prognosis patients with severely limited embryo cohorts, Day 3 (cleavage-stage) transfer remains a clinically recognized strategy to mitigate the risk of *in vitro* attrition[31-33]. Thus, SpermSeek reliably recovers high-quality gametes, actualizing the latent reproductive potential of these previously inaccessible samples.

Several limitations of the current study warrant consideration. First, strict ethical prohibitions against the uterine transfer of research-generated human embryos preclude the direct assessment of *in vivo* developmental trajectories and live-birth outcomes for these euploid Day 3 embryos. Second, while our findings functionally confirm the developmental competence of these rare NOA gametes up to the cleavage stage, their detailed transcriptomic and epigenetic landscapes remain to be fully mapped. Although defining these mechanisms is beyond the scope of the present study, future single-cell multi-omic profiling will be essential to unravel the precise molecular determinants—such as embryonic genome activation (EGA) dynamics—that govern extended *in vitro* progression. Finally, larger, multi-center prospective cohorts are required to validate rare-sperm recovery rates and standardize this platform across the diverse etiological and histopathological spectrum of NOA.

In summary, we establish an integrated AI-guided microfluidic platform capable of identifying and non-invasively recovering functionally competent gametes from a broad spectrum of NOA patients, including those for whom prior surgical extraction and prolonged pharmacological interventions yielded no sperm. This approach not only preserves sperm integrity to support ICSI fertilization and the subsequent development of euploid cleavage-stage embryos, but also provides compelling evidence that the clinical diagnosis of spermatogenic failure is far less definitive than traditionally presumed. Beyond NOA, this operator-independent system holds immediate translational potential for managing cryptozoospermia and severe oligozoospermia. By overcoming the physical constraints of conventional rare-cell retrieval—which relies on harsh centrifugation to concentrate samples for manual review—it offers a rapid, non-destructive, and scalable solution to standardize rare-gamete recovery. By bridging the ultra-low-latency microfluidics with reproductive biology, the SpermSeek platform extends the therapeutic horizon for patients unresponsive to conventional interventions and opens new frontiers for exploring the biological boundaries of human spermatogenesis.

## Materials and Methods

### 1. SpermSeek: A closed-loop, vision-activated microfluidic platform for non-destructive rare-sperm isolation

#### 1.1 System architecture and hardware synchronization

The SpermSeek platform was engineered as an integrated closed-loop system comprising a high-speed microscopic imaging module, a pressure-driven microfluidic interface, a continuous-flow piezoelectric droplet-printing unit, and a customized computing and control system. The imaging pathway utilized a custom-integrated high-speed CMOS vision module coupled with a 20× magnification objective (NA = 1.2). To capture transient sperm morphology without motion blur at a continuous volumetric throughput of 6 μL min⁻¹ (corresponding to a flow velocity of 23 mm s⁻¹), images were acquired at an operational frame rate of 180 fps. This specific rate was chosen to accommodate the 300–500 μs exposure time required for optimal phase-contrast illumination, although the camera hardware supports up to 330 fps at full resolution (4000 × 3700 pixels). To maximize data transfer efficiency and minimize computational overhead, real-time AI inference was executed on a targeted region of interest (ROI: 1000 × 3600 pixels) with a spatial resolution of 0.25 μm/pixel.

We developed a multi-threaded GPU-CPU heterogeneous computing framework. The CPU managed continuous image stream input/output (I/O) and the generation of physical actuation triggers, while a dedicated GPU (GeForce RTX 4090, NVIDIA) executed the deep-learning inference pipeline asynchronously. Inter-module coordination was managed through a purely software-defined synchronization pipeline. By establishing a direct, low-latency communication pathway between the GPU inference output and the CPU actuation module, this architecture bypassed the conventional reliance on complex, custom hardware logic (such as Field-Programmable Gate Arrays, FPGAs) typically required for sub-millisecond determinism. By minimizing buffering and latency-intensive OS-level scheduling delays, this software-centric design ensured that the sequential pipeline—linking image acquisition, tensor inference, and pneumatically-coupled piezoelectric actuation—supported tightly synchronized real-time operation of the closed-loop system.

#### 1.2 Real-time AI-based sperm identification

To balance real-time processing latency (∼3ms per frame inference budget) with high specificity against the complex cellular background of NOA semen, we deployed a highly optimized, single-stage object detection neural network based on the YOLOv8 architecture (specifically, the lightweight YOLOv8-Nano variant). To minimize computational overhead during high-frame-rate streaming, the model was tailored to process RGB image streams directly extracted from the high-speed camera.

The model was trained on an annotated dataset comprising approximately 10,000 microscopic images. To prevent overfitting and enhance robustness against dynamic flow artifacts, the training pipeline incorporated data augmentation, including synthetic motion blur, dynamic contrast adjustments, and the artificial superimposition of negative-class distractors (e.g., seminal plasma granularity, round cells, and leukocytes).

To evaluate the model’s performance on an independent hold-out test set (600 images), we employed a dual-metric approach. First, standard computer vision metrics were calculated using an Intersection over Union (IoU) threshold of 0.5 and strict class distinction. Second, to assess reliability under actual microfluidic actuation conditions, an expert-guided evaluation was conducted using pragmatically relaxed, clinically relevant criteria. Because the ∼20 nL droplet generated by the downstream piezoelectric actuator inherently tolerates minor spatial offsets, exact bounding box regression is unnecessary. Therefore, a detection was scored as a "true positive" if the target was contained anywhere within the predicted bounding box. Furthermore, because downstream clinical workflows inherently include a secondary morphological verification step, encapsulating representative ’sperm-like’ entities ensures the retrieval of actionable gametes. Thus, under this task-driven clinical metric, both typical sperm and sperm-like cells were counted as correct identifications. The resulting baseline algorithmic metrics and the clinically relevant performance outcomes (Precision, Recall, and False Positive Rates) are detailed in the Results section.

#### 1.3 Quantitative recovery validation via deterministic spiking

To benchmark the clinical recovery efficiency of SpermSeek, we established a deterministic spiking protocol using sperm-free seminal plasma prepared from healthy donors. To create this "zero-sperm" baseline, these healthy donor samples were pooled and subjected to three consecutive rounds of centrifugation (12000 × *g* for 10min). Confirmed by the absence of endogenous sperm in the resulting pellets, the pooled sample was established as a zero-sperm matrix.

To prepare standardized test samples with absolute target counts, we utilized the SpermSeek platform’s single-cell printing capability to "digitally" spike a known number of motile sperm into 1 mL aliquots of the verified sperm-free matrix. This approach allowed precise control of spiked sperm concentrations, creating groups of 10, 30, 100, 200, 400, 500, 800, and 1000 sperm mL⁻¹. For the standard clinical protocol (centrifugation-based control), spiked samples ranging from 30 to 1000 sperm mL⁻¹ were processed in triplicate (n = 3). To specifically validate SpermSeek’s performance at the extreme limits of NOA (the clinical ’blind spot’), experimental processing via the microfluidic platform focused on ultra-low concentration cohorts (spanning 10 to 400 sperm mL⁻¹ in triplicate, n = 3), alongside an evaluation at 800 sperm mL⁻¹ performed in duplicate (n = 2), evaluating its ability to rescue rare target cells that are inherently unrecoverable by standard means. For performance comparison, each spiked sample was divided into two equal groups. The control group was processed via the standard clinical protocol (centrifugation at 3000 × *g* for 15 min followed by manual pellet screening). The experimental group was processed through the SpermSeek autonomous identification and printing pipeline at a throughput of 0.36 mL h⁻¹ (corresponding to 6 μL min⁻¹). The number of recovered sperm in each group was quantified to evaluate the macroscopic recovery rate across the clinical concentration spectrum.

### 2. Microfluidic design for precise and stable single-sperm printing

#### 2.1 Microfluidic chip fabrication

The core microfluidic chip consists of a single-layer polydimethylsiloxane (PDMS) channel network designed for the precise manipulation and printing of individual sperm (Fig. 2a). The fluidic architecture is defined by two orthogonally intersecting pathways. The primary flow path sequentially comprises a semen reservoir (∼0.7 mL), a serpentine stabilization region, a high-aspect-ratio main channel (500 µm wide, 50 µm high), and a terminal waste outlet. The sample suspension is driven from the reservoir through the serpentine region, which imparts defined hydraulic resistance to stabilize the laminar flow profile before reaching the optical recognition and printing cross-junction. At this localized region, the horizontal channel (500 µm wide, 8 µm high) is orthogonally intersected by a vertically oriented printing channel (530 µm wide, 8 µm high), consisting of an upper printing buffer reservoir (∼25 µL) that feeds directly into the junction and terminates in a downward-facing micro-nozzle. Upon real-time target identification, a localized, transient pressure pulse (typically 2–8 ms) is actuated through the printing buffer reservoir, redirecting the identified sperm out of the horizontal streamline and downward through the nozzle to form a single-sperm encapsulated droplet.

The device was fabricated using a hybrid soft lithography approach. While the high-resolution microchannel networks were patterned on silicon wafers via standard photolithography[34, 35], the macroscopic fluid reservoirs (2 mm in height) exceeded standard photoresist limits. To integrate these disparate scales, custom 3D-printed reservoir molds were aligned and directly mounted onto the silicon master prior to casting PDMS prepolymer (SYLGARD™ 184, 10:1 w/w). Following thermal curing and demolding, the resulting monolithic PDMS replica was permanently bonded to a glass slide via oxygen plasma treatment. To suppress nonspecific cell adhesion, the microchannels were passivated with Pluronic F-127 as previously described[36], and conditioned with standard PBS prior to all assays.

#### 2.2 Printing dynamics and mechanical characterization

High-speed imaging was employed to characterize the microfluidic droplet-printing kinematics. A high-speed camera (Phantom, equipped with a 4× objective) was integrated with the SpermSeek platform to capture ejection dynamics and single-cell trajectories at the printing nozzle (Fig. 2a). Images were acquired at 6,000 fps with a 160-µs exposure time. To establish a kinematic baseline, the printing of Milli-Q water under various actuation pressures was first imaged to determine the initial ejection velocity, while the mean discharged volume per actuation was quantified gravimetrically (Fig. 2b). Subsequently, to define the operational phase boundary while decoupling bulk viscosity from seminal debris-induced perturbations, viscosity-matched aqueous glycerol-water surrogates (spanning the clinically relevant range up to 23 mPa·s) were evaluated across a combinatorial matrix of varying actuation pressures and defined pulse durations (2, 4, 6, and 8 ms). This multiparametric screening enabled the construction of a comprehensive operational phase diagram (Fig. 2d).

To quantify the transient mechanical loads exerted on sperm, we combined COMSOL Multiphysics (v6.1)-based simulations with experimentally constrained impact-load modeling. Corresponding to the experimental observations, the computational model evaluated two critical dynamic stages: (i) the rapid in-channel flow redirection at the printing junction, and (ii) the off-chip droplet impact onto a rigid collection substrate across practical dispensing heights (10–35 mm).

For the computational fluid dynamics (CFD) modeling, the transient in-channel velocity and pressure fields during flow redirection were reconstructed using the Laminar Flow interface in COMSOL, parameterized by experimentally calibrated pressure profiles. To isolate the effects of cellular volume while abstracting complex morphological variables, a rigid spherical surrogate (5 µm diameter, density 1050 kg m⁻³) was employed in the Particle Tracing module to evaluate the transient hydrodynamic loading. For the subsequent external droplet impact stage, local peak impact intensity and momentum-transfer equivalent impact loads were estimated using fluid-mechanics-derived models. The kinematic inputs for this external impact analysis—specifically, the ejection velocity and discharged volume—were defined using empirical values obtained from the high-speed imaging and gravimetric measurements (Fig. 2c).

### 3. Systematic profiling of rare sperm across the diverse spectrum of NOA

#### 3.1 Patient recruitment and clinical characterization

All human semen samples and corresponding clinical metadata were provided by Shenzhen Zhongshan Obstetrics & Gynecology Hospital. A defined cohort of 59 patients diagnosed with NOA was recruited for this study. The diagnosis of NOA was established based on the repeated absence of sperm in standard semen analyses following pelleting, combined with clinical evaluations to exclude obstructive etiologies. Comprehensive clinical histories—including etiological factors (e.g., genetic/chromosomal abnormalities such as AZF deletions, acquired injury), histopathological diagnoses (e.g., Sertoli cell–only syndrome, maturation arrest), and prior unsuccessful clinical interventions (e.g., pharmacological therapy, micro-TESE)—were obtained through a combination of institutional electronic medical records, clinical consultations, and validated patient-provided diagnostic reports to ensure accurate patient stratification.

#### 3.2 Semen processing and analytical microfluidic screening

Semen samples were collected via masturbation following a recommended abstinence period of 2–7 days and allowed to fully liquefy at room temperature, in accordance with the World Health Organization (WHO) Laboratory Manual for the Examination and Processing of Human Semen (6th Edition).

For a specific subset of samples presenting with dense cellular debris or hyper-viscosity—which posed a risk of microfluidic channel occlusion—a modified preprocessing step was implemented. These samples were subjected to centrifugation (3000 × *g* for 3–5 min). Given the established absence of sperm in the conventional diagnostic pellet of these NOA patients, we harvested the buoyant supernatant fraction rather than the cellular pellet. This fraction was subjected to the analytical screening workflow, targeting non-sedimenting, ultra-rare gametes while precipitating occlusion-causing detritus.

Following liquefaction or the aforementioned modified centrifugation step, to systematically map the prevalence and concentration of rare sperm across the NOA cohort, samples were processed using the analytical screening mode. In this configuration, the SpermSeek high-speed imaging module and YOLOv8 recognition software were integrated with a standard inverted microscope. Samples flowed through the microfluidic chip for real-time algorithmic identification, enumeration, and photographic logging, without physical droplet isolation.

#### 3.3 Morphological criteria and sperm quantification

To mitigate false-positive identifications in NOA samples, morphological criteria were established and enforced by a dual-expert verification protocol. A biological entity was classified as a "morphologically identifiable sperm" only upon joint confirmation by two nationally certified sperm analysis specialists (trained under WHO-standardized protocols, each with >7 years of clinical experience). Specifically, both experts had to verify that the entity exhibited the essential architecture of a human sperm: a discernible head structure coupled with an intact flagellar tail.

Any detected object causing disagreement between the two experts was excluded from the final count. Furthermore, ambiguous cellular debris or "sperm-like structures" lacking complete architectural integrity (e.g., isolated tail-like fragments or anucleate cell-like structures), as well as objects possessing atypically short tails or non-flagellar tail-like extensions, were excluded from the final quantitative analysis. Finally, the sperm concentration for each patient was determined by normalizing the filtered counts—derived from either the screening or recovery mode—to the initial volume of the native semen specimen.

### 4. Functional, epigenetic, and *in vivo* reproductive validation of SpermSeek-processed sperm

#### 4.1 Safety assessment of sperm function

To evaluate the basal functional and molecular safety of the SpermSeek microfluidic printing process, paired residual clinical semen samples from healthy donors (n = 16) were divided into unmanipulated controls and SpermSeek-processed groups. Sperm total motility (TM) and progressive motility (PM) were assessed using a computer-assisted semen analysis (CASA) system (Microptic). Intracellular reactive oxygen species (ROS) levels were quantified via flow cytometry using the DCFH-DA staining kit (BrayDe Biotech, Shenzhen, China). Sperm DNA integrity was evaluated by calculating the DNA fragmentation index (DFI) using the sperm chromatin structure assay kit (Anke Biotechnology, Hefei, China) coupled with flow cytometry.

#### 4.2 Whole-genome bisulfite sequencing (WGBS) and data analysis

Genomic DNA was extracted from paired sperm samples (n = 5 per group) using the MagPure Tissue & Blood DNA LQ kit (Magen) supplemented with DTT for membrane disruption. Bisulfite conversion was performed using the EZ DNA Methylation-Lightning Kit (Zymo Research), followed by WGBS library construction via the EpiArt® DNA Methylation Library Kit (Vazyme). Sequencing was conducted on an Illumina NovaSeq X Plus platform. Raw reads were filtered using fqtools and aligned to the human reference genome with Bismark. Differentially methylated regions (DMRs) were identified using the DSS package based on a beta-binomial distribution model, incorporating spatial smoothing.

#### 4.3 Mouse ICSI and multigenerational reproductive validation

All animal procedures were conducted in a specific pathogen-free (SPF) facility and approved by the Institutional Animal Care and Use Committee (IACUC) of GemPharmatech Co., Ltd. (Approval Nos. GPTAP032 for micromanipulation and GPTAP002 for husbandry/breeding). Wild-type C57BL/6JGpt male mice (≥10 weeks old) were utilized for sperm collection. Epididymal sperm were either processed through the microfluidic printing platform (intervention group) or utilized directly (control group).

Following standard hormonal superovulation of 3.5–4-week-old C57BL/6JGpt females, MII oocytes were collected and fertilized via intracytoplasmic sperm injection (ICSI) without additional chemical activation. For each group, 50 MII oocytes were injected with single immobilized sperm heads and cultured in KSOM medium. Embryos progressing to the 2-cell stage were surgically transferred into the oviducts of pseudopregnant ICR/Gpt recipients (n = 2 per experimental group) under Ketamine/Xylazine anesthesia.

To evaluate transgenerational safety, the resulting F1 progeny were monitored for developmental milestones, body weight, and neonatal survival. Upon reaching sexual maturity (∼9 weeks of age), F1 mice were crossed (1:1 ratio) with naturally bred wild-type C57BL/6JGpt mice to generate the F2 cohort.

### 5. Embryogenic potential and genomic stability of rare sperm in NOA

#### 5.1 Clinical cohort and SpermSeek recovery

To isolate functional gametes for downstream evaluation, native semen samples from the validation cohort (n = 26 NOA patients) were processed following the clinical protocols detailed in Section 3.2. Prepared samples were loaded into the fully integrated SpermSeek instrument. Upon real-time algorithmic identification of a target sperm, the system actuated a calibrated pneumatic pulse (1800 mbar, 4 ms pulse duration) to encapsulate and orthogonally eject the cell into a collection dish.

#### 5.2 Research-only ICSI and Time-Lapse Embryo Culture

To evaluate the embryogenic competence of SpermSeek-recovered gametes, research-only ICSI was conducted. Anonymized mature (MII) oocytes were obtained from healthy donors who provided written informed consent. Resulting embryos were generated for research purposes to assess the biological boundaries of NOA spermatogenesis; these embryos were legally and ethically prohibited from clinical uterine transfer.

Following microfluidic printing into collection dishes, individual target spermatozoa were microscopically localized and immediately cryopreserved utilizing standard single-sperm vitrification protocols. Following rapid thawing and removal of cryoprotectants, structurally intact sperm were selected. Single-sperm injection into donor MII oocytes was performed using standard micromanipulation procedures. Injected oocytes were subsequently cultured in Quinn’s Medium (SAGE, USA), and monitored in time-lapse incubators: Embryoscope (Vitrolife, Gothenburg, Sweden) or GERI (Genea Biomedx, Sydney, Australia) from Day 1 to Day 7.

#### 5.3 Whole-Embryo Lysis and Preimplantation Genetic Testing (PGT-A)

To evaluate genomic stability of the paternal contribution, all developing embryos (n=18) underwent preimplantation genetic testing for aneuploidy (PGT-A). As the embryos obtained in this research project are prohibited from being used for clinical transfer, trophectoderm biopsy was omitted; instead, whole embryos were lysed to maximize DNA yield. Whole genome amplification (WGA) and library preparation were performed using the ChromInst™ Universal Library Preparation Kit, followed by sequencing on a BGI DNBSEQ T7 platform (approximately 3 million raw reads per sample). Sequencing data were aligned to GRCh37/hg19, and copy number analysis was conducted using 400 kb sliding windows (200 kb step size). Segments with log₂ ratio ≥ ±0.25 were identified by circular binary segmentation (CBS) to detect whole chromosome aneuploidies, mosaicism (≥ 10 Mb, >30% level), and major segmental abnormalities (≥4 Mb).

### 6. Statistical analysis

Statistical analyses were performed using GraphPad Prism 10.1.2 (GraphPad Software, San Diego, CA, USA) and SPSS version 23 (IBM, Chicago, IL, USA). For categorical variables, differences between groups were assessed using Fisher’s Exact Test. For continuous variables, comparisons between independent groups were conducted using the Mann–Whitney U test, while paired measurements from the same original sample were analyzed using the Wilcoxon signed-rank test. All tests were two-sided, and a P value < 0.05 was considered statistically significant.

### 7. Ethics declarations

This study was approved by the Ethics Committee of Shenzhen Zhongshan Obstetrics & Gynecology Hospital and has been under ongoing ethical oversight (Approval Nos: SZZSECHU-2023004, SZZSECHU-2025081). Written informed consent was obtained from all participants.

## Data Availability

All data produced in the present study are available upon reasonable request to the authors

## Acknowlegements

The initial conceptualization, full engineering architecture, algorithm training and software of the SpermSeek platform were exclusively funded by Zhuhai Dalue Technology, Ltd (DalueTech). We thank Kang Li for his contribution to the low-level electrical control and microcontroller interface design of the platform. The subsequent clinical application—including the fabrication of device replicas and consumables, murine experiments, epigenomic profiling, and embryological evaluations—was supported by Guangdong Basic and Applied Basic Research Foundation (Grant No. 2026A1515011720); Shenzhen Science and Technology Innovation Commission (JCYJ20241202123732042); Shenzhen Municipal Medical Research Special Fund (Grant No. D250402002).

## Author Contributions

H.C. conceived the original concept, directed the entire study, engineered the core device architecture, secured funding, and wrote the manuscript. P.C. designed and supervised the downstream biological and pre-clinical validation pipelines, and analyzed the associated data. W.X. led the clinical translation phase, directed patient and oocyte volunteer recruitment, and interpreted clinical endpoints. L.W. performed the microfluidic chip simulations and validations, analyzed experimental data, and generated the manuscript figures. M.S. facilitated essential institutional ethics coordination for embryo experiments and assisted in clinical logistics. X.L. and R.S. validated the system and conducted the core sperm detection and recovery assays, processed clinical samples. S.G., J.L., and W.Z. collaboratively developed the device hardware, control software, AI algorithms, and executed system integrations. M.M., C.H., and S.X. contributed to ethical approval, clinical governance, safety and efficacy assessment, and coordination of oocyte volunteer recruitment. Q.S., H.X.Z., and L.Y. performed reproductive procedures, embryo validation, safety assays, and sperm functional analyses. Y.X. fabricated the 3D-printed microfluidic molds. C.C. contributed to early manuscript drafting, figure preparation, and literature review. F.X., H.Z.Z., and X.W. assisted with ethics implementation, embryo-validation support, and recruitment execution.

## Competing Interests

H.C. is the founder and holds equity in Zhuhai Dalue Technology, Ltd (DalueTech), a university-affiliated spin-off company. W.Z., X.L., S.G., J.L., K.L., and R.S. are employees of DalueTech. The core hardware, software, and microfluidic architectures of the SpermSeek platform were developed at DalueTech, which owns the related intellectual property and pending patents. The remaining authors declare no competing interests.

